# How “Micro” is Microperimetry? Characterizing the Effect of Fundus Tracking on the Psychometric Function

**DOI:** 10.64898/2026.03.25.26349170

**Authors:** Theresa Lipsky, Clara Ehrenzeller, Georg Ansari, Kristina Pfau, Wolf Harmening, Zhichao Wu, Maximilian Pfau

## Abstract

**Purpose:** To quantify whether fundus tracking in microperimetry improves psychometric parameter estimation (*in vivo* demonstration of improved stimulus-delivery precision), and to derive a psychometrically grounded criterion intensity for suprathreshold (defect-mapping) microperimetry.

**Methods:** Twenty-five healthy volunteers underwent MAIA2-microperimetry at five loci: three outside and two inside the blind spot. Frequency-of-seeing (FoS) functions were measured in four blocks (2 tracking on; 2 tracking off). FoS-data were fit using cumulative-Gaussian psychometric functions estimating sensitivity parameters. Mixed-effect models assessed tracking effects, and posterior simulations defined the optimal criterion intensity for separating “seeing” from “non-seeing” loci.

**Results:** Tracking had little effect on threshold estimates at loci outside the blind spot, but lowered threshold estimates within the blind spot (posterior median difference PMD [95% CrI] of −1.46 dB [−2.30, −0.62] at locus 4, and −1.02 dB [−1.94, −0.08] at locus 5). Tracking was associated with steeper psychometric slope parameters at loci 1–3 (PMD of −0.14 dB [−0.29, 0.01], −0.27 dB [−0.43, −0.12], and −0.22 dB [−0.40, −0.04]). Without tracking, false-positive responses were more frequent when fixation shifts displaced stimuli toward the “seeing” retina. Simulation-based analysis identified 13 dB as nominally optimal criterion for suprathreshold microperimetry (Youden index: 0.76 [0.74, 0.79], comparable to 10 dB (0.74 [0.72, 0.76]).

**Conclusions:** Even in healthy volunteers with stable fixation, fundus tracking measurably reduced sensitivity estimates at “non-seeing” loci and sharpened FoS curves in the “seeing” retina. A criterion intensity of 10 to 13 dB is a defensible choice for separating “seeing” and “non-seeing” retina in suprathreshold (defect-mapping) perimetry paradigms.

## INTRODUCTION

Microperimetry (fundus-controlled perimetry) has matured from a research technique into a standard clinical trial tool for macular visual field testing.^1^ By visualizing the fundus with a scanning laser ophthalmoscope or a fundus camera in real time, stimuli can be presented at predefined retinal locations, even in patients with unstable fixation.^1^ Especially in geographic atrophy (GA) secondary to age-related macular degeneration (AMD), microperimetry is becoming increasingly important as it enables tracking disease progression in eyes showing no change in best-corrected visual acuity due to foveal sparing or early foveal involvement.^2,3^

To date, it is unclear how accurately microperimetry devices can track and present stimuli at the intended retinal location, given the complexity of fixational eye movements, including microsaccades, drift, and torsional motion.^4,5^ Clinical observations provide face validity, because test points within areas of GA, especially centrally located within lesions, typically show no response, whereas points just outside the GA boundary exhibit a steep functional increase within the outer ∼500 µm junctional zone. This mirrors the pattern of photoreceptor loss seen in histopathology.^2,6^ Moreover, localized optical coherence tomography data can be used to predict function with high accuracy indicating that stimuli are, *on average,* presented where intended.^7–9^

However, these demonstrations of face validity rely on aggregates across stimulus presentations and do not directly answer a key psychophysical question: how precisely is each individual stimulus placed at the intended retinal location? This question becomes critical in junctional zones with steep sensitivity gradients, where even sub-degree placement errors or small fixation excursions can flip a response from “seen” to “not seen.” Although stimulus-placement accuracy has been examined previously for a modified MP1 microperimeter, those findings are difficult to interpret in the present context owing to the device-specific modifications employed and the obsolescence of the MP1 platform.^10^

Although GA provides a clinically relevant setting for understanding the effect of tracking on the psychometric function, it is not an ideal experimental model for isolating stimulus-delivery precision. Areas of atrophy do not uniformly represent absolute scotomata, but can still yield residual detections, particularly at loci with a residual outer nuclear layer.^11^ The physiologic blind spot provides a useful complementary model for studying stimulus-delivery precision. The optic nerve head constitutes an anatomically defined deep scotoma with a comparatively sharp boundary and, unlike GA, is not confounded by variable residual photoreceptor structure.^12^

Knowledge of the psychometric function close to scotomata boundaries is particularly important for suprathreshold testing paradigms that aim to separate “seeing” from “non-seeing” retina with a single presentation.^8,12^ Ideally, the fixed *criterion intensity* for suprathreshold perimetry should be justifiable based on both clinical heuristics,^13^ as well as the psychometric function.

Fixed-stimulus testing, analyzed through frequency-of-seeing (FoS) modeling, provides the natural framework for defining psychometric cut-offs between “seeing” and “non-seeing” retina.^14^ FoS data under mesopic conditions are available (mirroring the MAIA2 test conditions), but remain limited and are absent for the MAIA2 device itself.^14^

These gaps motivate a closer examination of how “micro” microperimetry truly is, using prospectively acquired FoS data with the MAIA device, with and without tracking, in healthy volunteers at three loci outside the physiologic blind spot and two loci within it, thereby leveraging the optic nerve head as a natural scotoma. Specifically, we asked: (i) Does fundus tracking provide stimulus-placement precision beyond natural fixation stability in young observers, thereby sharpening FoS threshold and slope parameter estimates? (ii) Does fixation-related stimulus misplacement in the absence of tracking systematically give rise to false-positive and false-negative responses at “non-seeing” and “seeing” loci, respectively? and (iii) Do these psychometric data support the current criterion intensity for defect-mapping microperimetry?

## METHODS

### Participants

The study was approved by the Ethics Committee for Northwestern and Central Switzerland (EKNZ) and conducted in accordance with the Declaration of Helsinki. All participants received written and verbal information about the procedures and provided written informed consent prior to participation.

Eligible participants were ≥18 years old and had no history of ocular surgery that, in the investigator’s judgement, could affect visual function outcomes. Surgeries permitted for inclusion were uncomplicated cataract extraction, YAG laser capsulotomy, and laser retinopexy.

### Clinical Examination

Participants underwent autorefraction, followed by best-corrected visual acuity testing. Retinal structure was assessed with spectral-domain optical coherence tomography (SD-OCT; Heidelberg Spectralis OCT2, Heidelberg Engineering, Heidelberg, Germany), including (1) a macular volume scan (30° × 25°, 121 B-scans, automatic real-time averaging = 25) and (2) an optic nerve head scan using the device’s preset Bruch’s membrane opening (BMO) protocol.

### Test Grid Design

The experiment was designed to use the optic nerve head as a natural deep scotoma with a sharp boundary.^12^ Using the OCT volume, an experienced grader identified the B-scan passing through the vertical midline of the optic disc and manually labeled the temporal Bruch’s membrane opening on that B-scan and on the co-acquired Spectralis infrared (IR) reference image.

A MAIA microperimetry examination was then initiated via the Open Perimetry Interface (OPI).^15,16^ At test start, OPI provides a “MAIA template IR” image, which served as the retinal tracking reference for the entire examination. The labeled Spectralis IR image was registered to the MAIA template IR image in ImageJ using a landmark-based transform (Landmark Correspondences). A custom ImageJ^17^ plugin then generated MAIA stimulus coordinates for five test locations positioned relative to the Bruch’s membrane opening border: three locations outside the Bruch’s membrane opening (+0.25°, +0.75°, +1.25°) and two locations inside the Bruch’s membrane opening (−0.25°, −0.75°). Offsets were chosen so that the Goldmann III target (0.43° diameter; 0.215° radius) did not fall exactly on the anatomical boundary.

### Psychophysical Experiment

Sensitivity at each location was first estimated using Zippy Estimation by Sequential Testing (ZEST).^18^ This estimate defined the stimulus range for frequency-of-seeing (FoS) measurements: FoS experiments spanned a total of seven intensity levels centered on the ZEST estimate and spanning ±3 dB (i.e., 1 dB steps). If the targeted range included steps below 0 dB, the FoS range was truncated to sensitivities of 0 dB or more.

FoS functions were then measured using a method of constant stimuli in a total of four blocks: two with retinal tracking enabled (blocks 1 and 3) and two with tracking disabled (blocks 2 and 4). The block order was randomized and participants were masked to the tracking condition (Figure 1). For each location, 15 presentations per intensity set were delivered per block (i.e., 15 × 7 = 105 presentations per location and block; slightly fewer for locations within Bruch’s membrane opening due to the floor of the sensitivity range at 0 dB).

**Figure 1.**
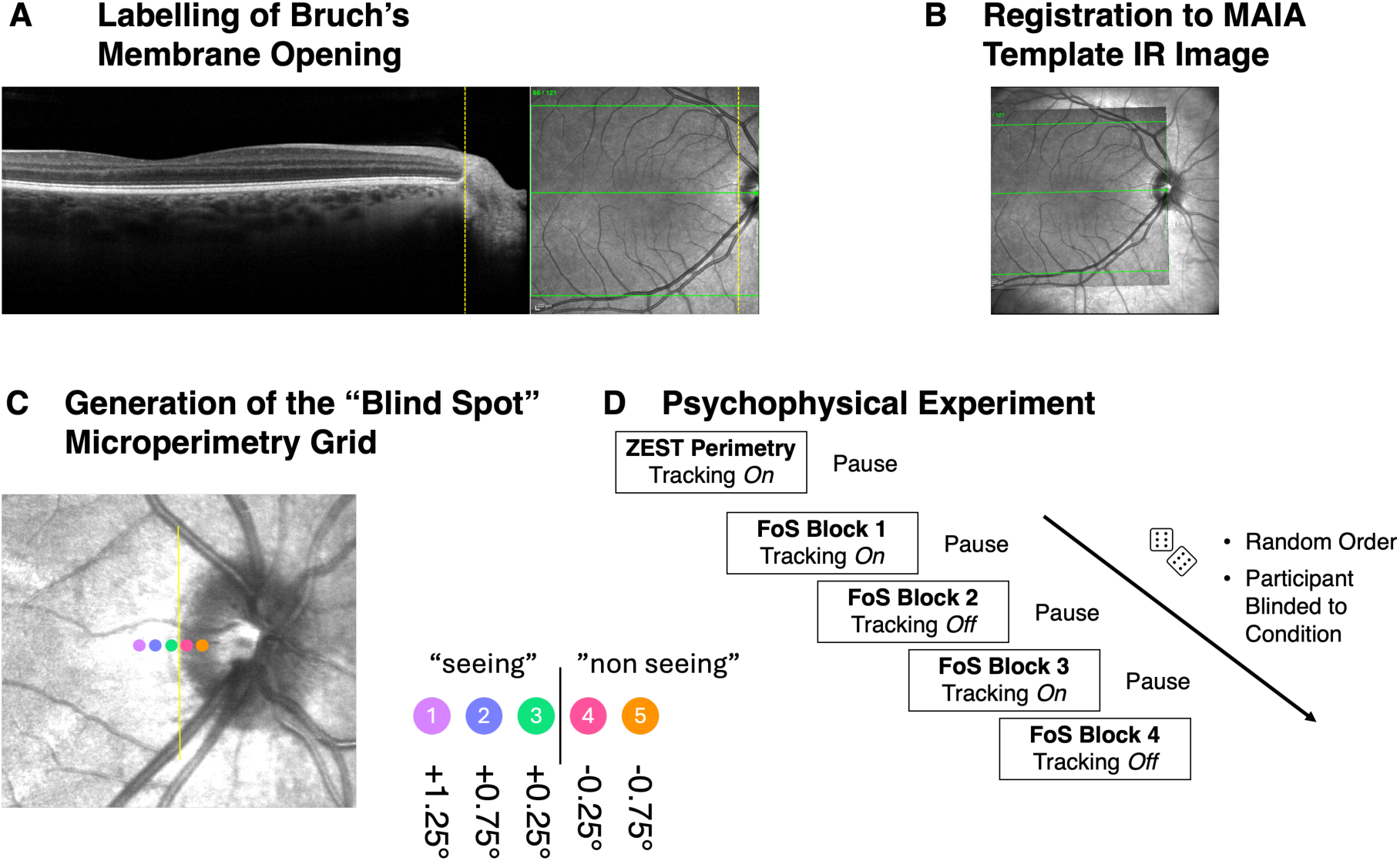
Experimental Design. The optic disc was used as a natural scotoma. **(A)** First, an optical coherence tomography scan was used to identify the temporal Burch’s membrane opening of the optic disc, and the label was transferred to the co-acquired infrared reflectance (IR) image (yellow dashed line). **(B)** The IR image and label were then registered to the MAIA device’s IR image from the start of the exam. **(C)** The testing grid comprised 5 locations with three locations outside of Burch’s membrane opening (+1.25 deg, +0.75 deg, and +0.25 deg) and two locations within Burch’s membrane opening (-0.25 deg, -0.75 deg). The marker size approximates the Goldmann III stimulus size. **(D)** The actual psychophysical experiment consisted of an initial perimetry test to define the range of sensitivity for each locus, followed by four blocks with frequency-of-seeing (FoS) testing. These were ordered randomly, and the participants were blinded to the condition (tracking enabled or disabled).

### Statistical Analysis

Participant and trial-level data are publicly available (*Zenodo repository to be shared upon acceptance*). All analyses were performed in R. Frequency-of-seeing (FoS) data were modeled for each subject, locus, and condition using a cumulative Gaussian psychometric function. The probability of reporting a stimulus as seen was expressed as

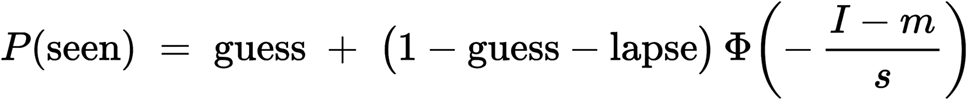

where *m* denotes the threshold corresponding to the midpoint of the function, *s* governs the slope parameter of the transition region, *I* represents the intensity in decibel (dB), *guess* represents the lower asymptote (false-positive rate), *lapse* represents the upper asymptote (false-negative rate), and Φ is the standard normal cumulative distribution function. Model fitting was performed within a Bayesian framework to allow direct propagation of uncertainty in parameter estimates (Figure 2).^19^

**Figure 2.**
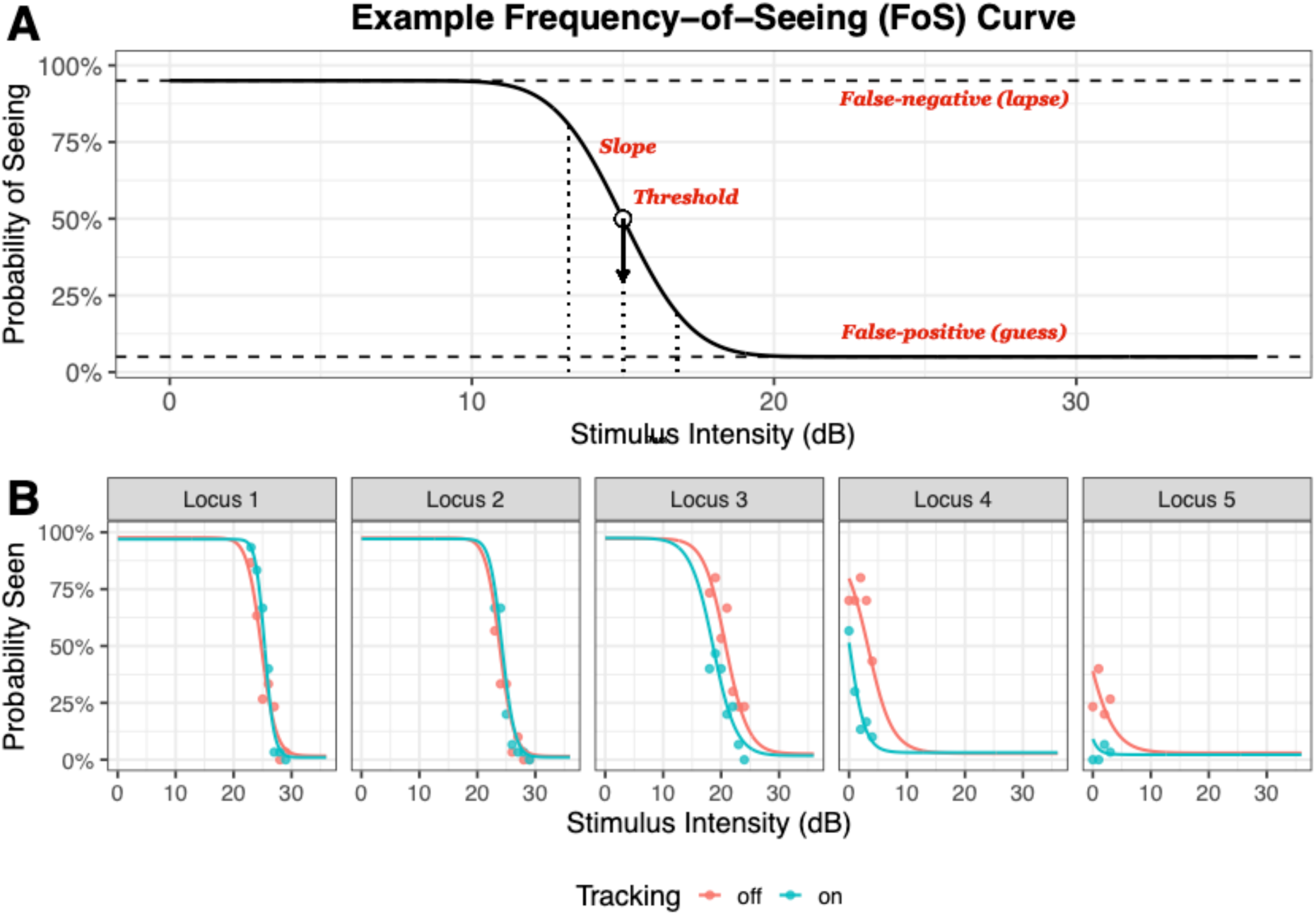
Frequency-of-seeing (FoS) psychometric function and example individual data. The top panel (**A**) shows a schematic frequency-of-seeing curve illustrating the main psychometric parameters. The probability of reporting a stimulus as seen is plotted as a function of stimulus intensity. The threshold (m) corresponds to the stimulus level at which the response probability reaches 50% after accounting for false-positive (guess) and false-negative (lapse) response rates. The slope parameter (s) describes the steepness of the transition from “non-seeing” to “seeing” around threshold. Dashed horizontal lines indicate the lower and upper asymptotes defined by guess and lapse rates. The bottom panel (**B**) shows example data from a single study participant showing fitted FoS curves at five retinal loci (columns). Points represent observed response probabilities at each stimulus intensity, and solid lines show the fitted psychometric functions. Data are shown separately for tracking-off (red) and tracking-on (blue) conditions. For loci 4 and 5 (targeting the “non-seeing” Bruch’s membrane opening), tracking results in a reduced rate of responses as anticipated.

Test–retest reliability was assessed separately for tracking-on and tracking-off conditions by comparing repeated blocks within each condition (tracking-on: FOS1 vs FOS3; tracking-off: FOS2 vs FOS4). Reliability was evaluated for threshold, slope parameter, guess, and lapse using Bland–Altman analyses implemented with *SimplyAgree*.^20^

The primary analysis of tracking effects on psychometric parameters was performed using a joint Bayesian multivariate model with threshold and slope parameter as correlated outcomes. For this analysis, first-stage threshold and slope parameter estimates were reshaped into a wide data set containing one row per subject, locus, block, and tracking condition. Threshold and slope parameter were modeled jointly as Gaussian outcomes with fixed effects for tracking, locus, and their interaction, and with shared subject-level random intercepts and random slopes for tracking. Outcome-specific inverse-variance weights, defined as the inverse squared standard errors from the first-stage psychometric fits, were used to account for differing precision of parameter estimates. Because different weights were used for threshold and slope parameter, residual correlations between outcomes were not estimated; however, cross-outcome associations at the subject level were retained through shared random effects. Posterior summaries were reported as posterior medians, 95% credible intervals (CrI), and probabilities of direction (pd). Population-level marginal means under tracking-off and locus-specific tracking effects (tracking-on minus tracking-off) were derived from posterior predictions.

As sensitivity analyses, threshold and slope parameter were also modeled separately using analogous Bayesian mixed-effects models fitted across all loci. In addition, for direct comparison with more conventional approaches, frequentist linear mixed-effects models were fitted separately for each locus using lme4, with tracking as a fixed effect and subject-specific random intercepts and slopes.

To examine fixation-related sources of likely false-positive and false-negative responses, fixation measurements were mapped to the timing of stimulus presentation. The fixation measurements obtained from the MAIA2 device via OPI were saved at a frequency of 1 per 642 ms (i.e., less than the device’s tracking frequency). As a result, the median absolute difference between fuzzy-matched stimulus presentation and fixation measurement pairs was 157 ms with a maximum difference of 341 ms.

Bayesian logistic mixed-effects models were fitted jointly across loci. For likely false-positive responses, the model included negative horizontal displacement (corresponding to displacement of the stimulus toward the “seeing” retina), tracking condition, locus, and all interactions among these terms, with subject-specific random intercepts. For likely false-negative responses, an analogous model was fitted using positive horizontal displacement (corresponding to displacement of the stimulus toward the optic nerve head), again including tracking, locus, and their interactions, with subject-specific random intercepts. Locus-specific odds ratios, 95% credible intervals, and probabilities of direction were derived from posterior linear predictors. As sensitivity analyses, frequentist per-locus logistic mixed-effects models were also fitted using glmer.

Optimal suprathreshold stimulus intensity for defect-mapping microperimetry was determined from operating characteristics derived from psychometric functions fitted under tracked conditions only, reflecting the clinically relevant examination setting. Posterior predictive simulations were used to propagate uncertainty arising from psychometric parameter estimation and binomial response variability at the trial level. True-positive and false-positive rates were estimated across the full range of stimulus intensities, and the criterion maximizing Youden’s index was identified, with uncertainty summarized using posterior credible intervals. As a sensitivity analysis, the same procedure was repeated using only locus 1 as the “seeing” locus and locus 5 as the “non-seeing “locus, to minimize potential confounding from peripapillary atrophy.

## RESULTS

### Demographics

A total of 25 healthy volunteers (median age [Q1, Q3]: 27.0 years [25.1, 30.1]; 12 females, 13 males) were enrolled. All participants exhibited normal ocular findings on slit-lamp examination and spectral-domain optical coherence tomography and had best-corrected visual acuity of 20/16 Snellen equivalent or better (median [Q1, Q3] of -0.1 logMAR [-0.1, - 0.1]).

The dataset comprised 500 independently measured frequency-of-seeing functions. The acquisition of a block took on average (median [Q1, Q3]) 21.3 min [19.7, 23.5] with tracking off and 23.3 min [20.9, 25.3] with tracking on.

### Locus-specific psychometric parameters and test–retest reliability

Absolute psychometric parameters differed systematically across loci, consistent with their retinal location relative to the blind spot (Table 1). Under tracking-off conditions, threshold estimates were highest at loci 1–3, with posterior medians of 25.19 dB (95% CrI: 23.79–26.54), 23.46 dB (22.05–24.77), and 20.12 dB (18.71–21.49), respectively. Thresholds declined sharply at loci 4 and 5, with posterior medians of 6.17 dB (4.69–7.59) at locus 4 and 0.64 dB (−0.89 to 2.03) at locus 5, approaching the measurement floor. Slope parameter estimates likewise varied across loci. Under tracking-off conditions, posterior median slope parameter estimates were 1.62 (1.43–1.80) at locus 1, 1.89 (1.70–2.08) at locus 2, 2.40 (2.19–2.59) at locus 3, 2.57 (2.36–2.79) at locus 4, and 2.00 (1.80–2.21) at locus 5.

**Table 1.**
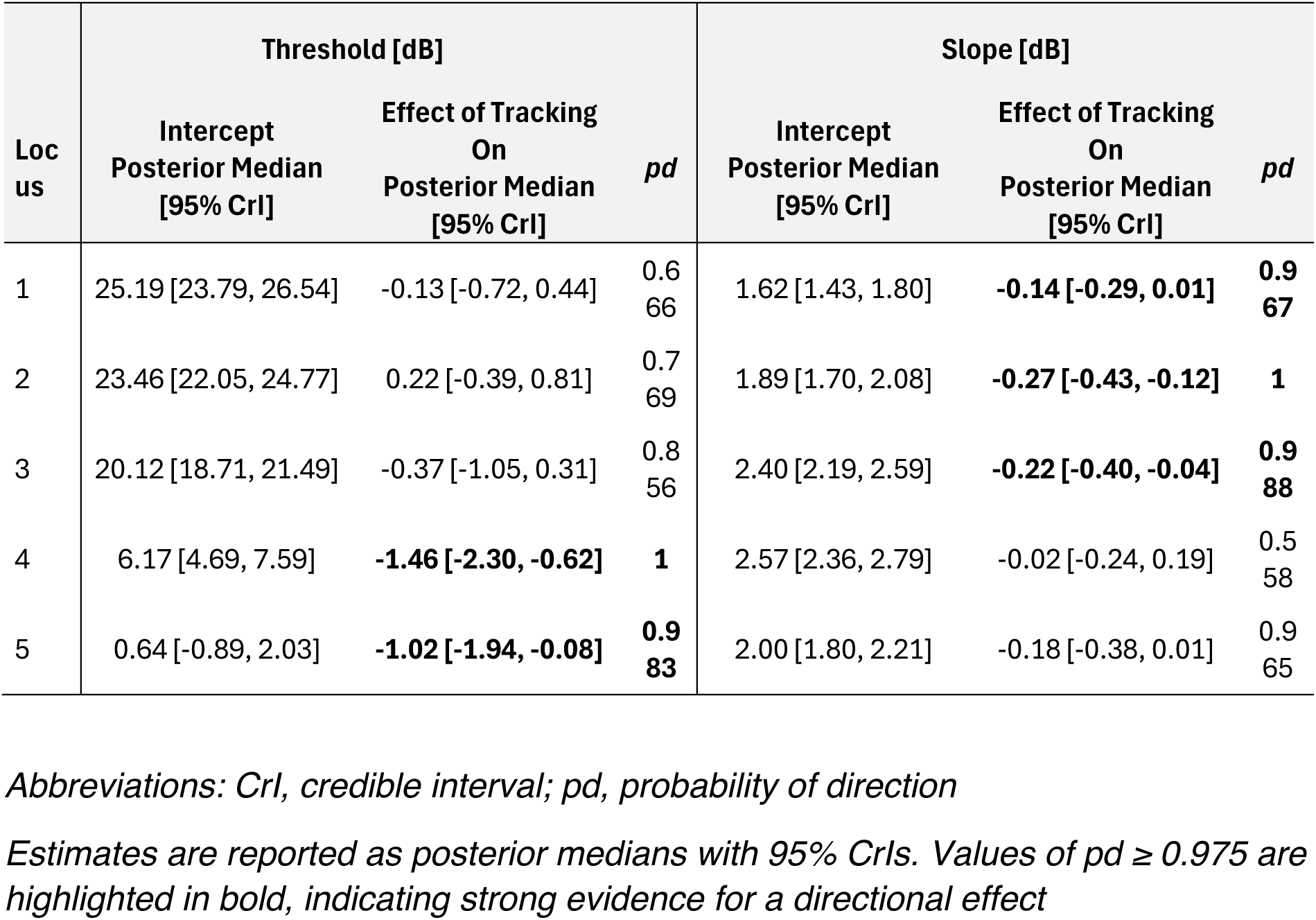
Estimates of the Joint Bayesian Model for Threshold and Slope Parameter Across Loci for Tracking On and Off.

Test–retest reliability was assessed separately for tracking-on and tracking-off conditions by comparing repeated blocks within each condition (blocks 1 vs 3 and blocks 2 vs 4, respectively). Bland–Altman analyses showed comparable repeatability with and without tracking across all psychometric parameters (Supplementary Figure S1; Supplementary Table S1). Threshold estimates (m) showed small biases and limits of agreement of approximately ±3 dB in both conditions, while slope parameter estimates (s) showed minimal bias and limits of agreement of approximately ±2 dB. Guess and lapse rates were highly stable, with near-zero biases and narrow limits of agreement irrespective of tracking status.

### Effect of fundus tracking on the psychometric function

Bayesian mixed-effects models were used to estimate the effect of retinal tracking on threshold and slope parameter at each test locus (Table 1; Figure 3). Across loci 1–3, located outside the blind spot, the effect of tracking on threshold was small and uncertain. Posterior median differences were −0.13 dB (95% CrI: −0.72 to 0.44; pd = 0.666) at locus 1, 0.22 dB (−0.39 to 0.81; pd = 0.769) at locus 2, and −0.37 dB (−1.05 to 0.31; pd = 0.856) at locus 3. Thus, for these loci, the posterior distributions were centered close to zero and the credible intervals overlapped zero.

**Figure 3.**
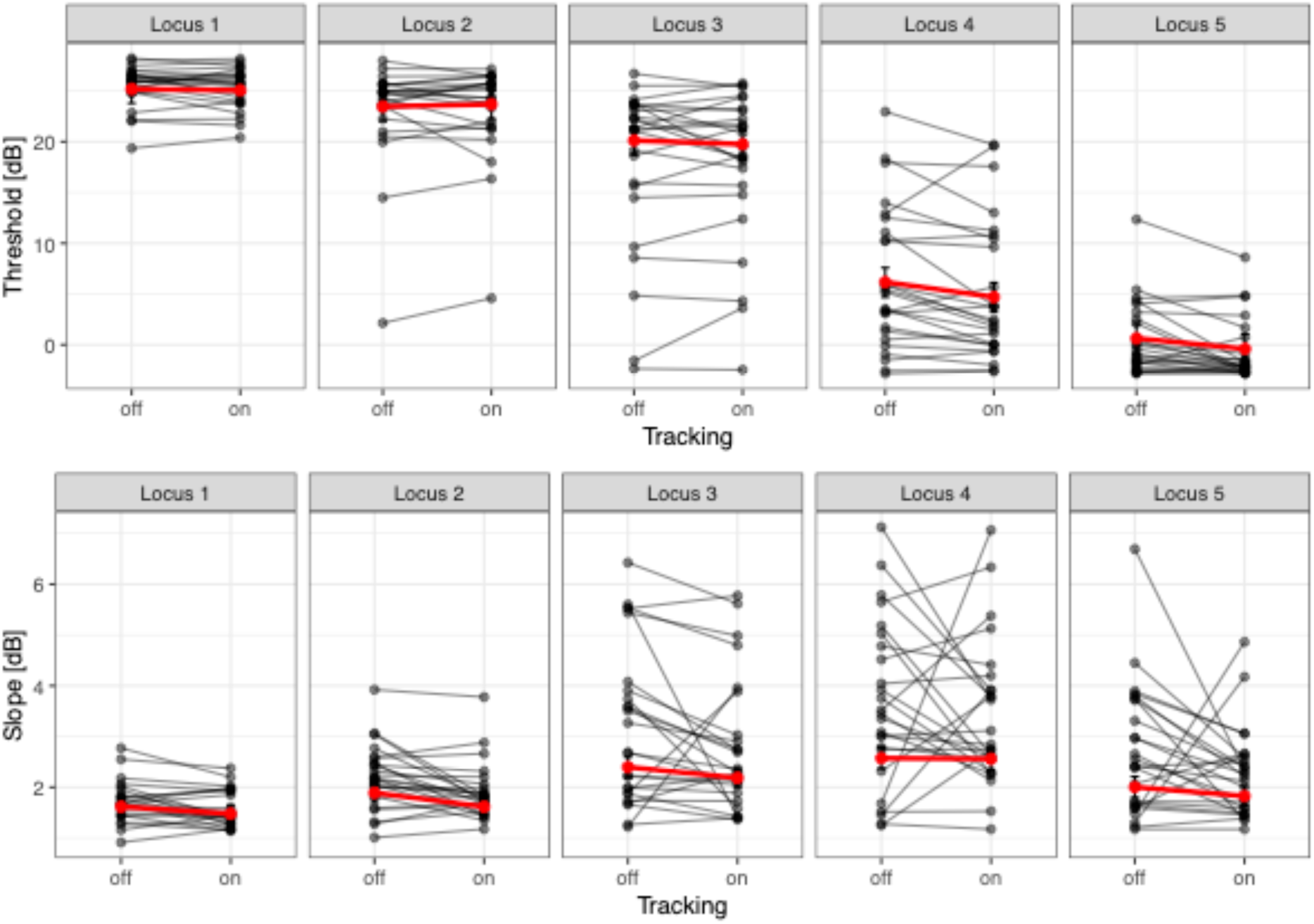
Effect of tracking on the psychometric function. Threshold (m, top row) and slope parameter (s, bottom row) estimates are shown for five retinal loci (columns). Gray points and connecting lines represent within-subject averages for tracking-off and tracking-on conditions. Red lines indicate the fixed-effect estimates from linear mixed-effects models. Across loci, tracking-related differences were generally small and comparable to test–retest variability. Modest reductions in threshold with tracking were observed at loci closer to the blind spot, while slope parameter estimates showed subtle, locus-dependent decreases with tracking enabled.

In contrast, at the two loci located within the blind spot, thresholds were lower when tracking was enabled. At locus 4, the posterior median difference was −1.46 dB (95% CrI: −2.30 to −0.62; pd = 1.000), and at locus 5 it was −1.02 dB (−1.94 to −0.08; pd = 0.983). These results indicate strong directional evidence for a negative effect of tracking on threshold at both blind-spot loci.

For slope parameter, tracking effects were more spatially heterogeneous. At loci 1–3, tracking was associated with steeper slope parameter estimates, with posterior median differences of −0.14 (95% CrI: −0.29 to 0.01; pd = 0.967) at locus 1, −0.27 (−0.43 to −0.12; pd = 1.000) at locus 2, and −0.22 (−0.40 to −0.04; pd = 0.988) at locus 3. By contrast, within the blind spot, there was little evidence that tracking altered slope parameter. Posterior median differences were −0.02 (−0.24 to 0.19; pd = 0.558) at locus 4 and −0.18 (−0.38 to 0.01; pd = 0.965) at locus 5, indicating no clear effect at locus 4 and only weak-to-moderate directional evidence at locus 5.

Consistent results – both for the effect of tracking on threshold and slope parameter – were obtained across all model specifications, including separate Bayesian models for threshold and slope parameter fitted across all loci as well as frequentist linear mixed-effects models fitted per locus (Supplementary Tables S2–S4).

### Identifying the casus of false-negative and -positive responses in the absence of tracking

To determine whether fixation position at or near stimulus presentation contributed to likely *false-positive responses*, we analyzed participant responses for stimuli with a low predicted probability of being seen (< 0.1, based on psychometric curve fits under tracked conditions). Across loci, responses obtained without tracking were consistently more likely to be reported as seen, with odds ratios ranging from 2.15 to 3.21 and strong directional evidence (pd ≥ 0.987 at all loci; Figure 4 and Supplementary Tables S5 and S6).

**Figure 4.**
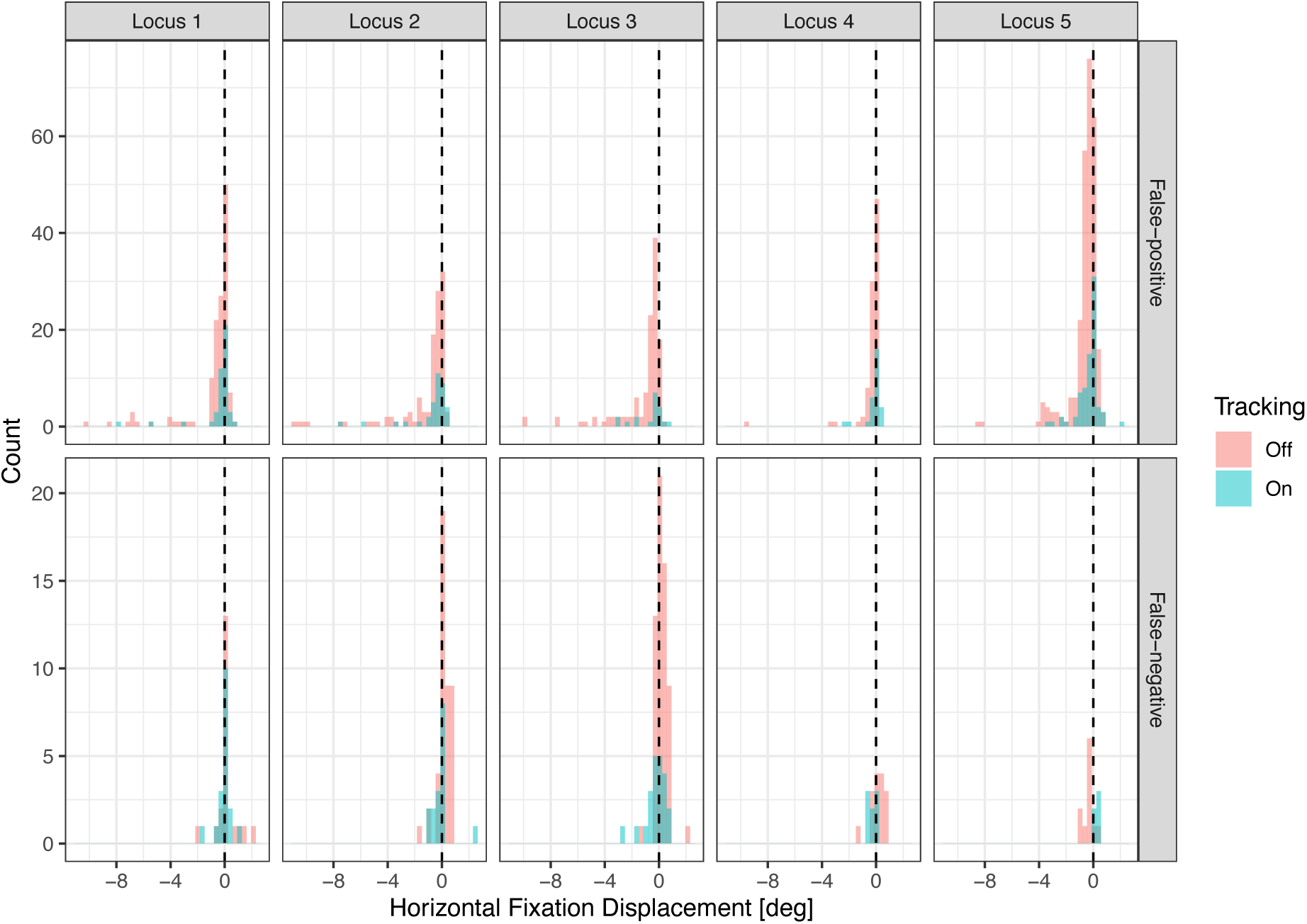
Spatial distribution of fixation at the time of likely false-positive and false-negative responses. Histograms show the horizontal eccentricity of fixation, evaluated at or near the time of stimulus presentation. Data are shown separately for five test loci (columns) and response type (rows: false-positive responses, top; false-negative responses, bottom). False-positive responses were defined as responses with a low predicted probability of being seen (< 0.1), whereas false-negative responses were defined as missed stimuli despite a high predicted probability of being seen (> 0.9). Within each panel, distributions are shown for tracking-off and tracking-on conditions. In the absence of tracking (red histograms), false-positive responses were associated with systematic horizontal fixation shifts resulting in a foveal-ward displacement of the stimulus (especially evident for locus 5 within the blind spot). Conversely, false-negative responses—particularly at locus 3 (just outside the blind spot)—were associated with fixation shifts in the opposite direction, leading to stimulus presentations displaced toward the optic nerve head. These patterns were markedly reduced under tracking-on conditions (green histograms).

A negative horizontal displacement (i.e., stimulus displacement toward the “seeing” retina) showed modest and heterogeneous effects across loci under tracking-on conditions: An increase in false-positive responses was evident at loci 2 and 5 (OR = 1.99 [1.00, 4.23], pd = 0.975; OR = 1.69 [1.06, 2.74], pd = 0.985), effects at other loci were smaller and more uncertain.

But, the interaction between a negative shift and tracking off showed strong evidence at selected loci, most notably at locus 3 (OR = 3.62 [1.24, 10.69], pd = 0.992) and locus 5 (OR = 2.90 [1.63, 5.26], pd = 1.000), indicating that fixation displacement substantially increased the likelihood of false-positive responses in the absence of tracking.

Conversely, for stimuli expected to be seen with high probability (> 0.9), fixation shifts displacing the stimuli toward the optic nerve head under tracking-off conditions were associated with a reduced probability of reporting the stimulus as seen, corresponding to an increased occurrence of *false-negative responses* (Figure 4 and Supplementary Tables S7 and S8). While main effects of horizontal displacement and tracking alone were generally small and uncertain, strong evidence for interaction effects was observed at loci 2–4.

Specifically, the odds of reporting a stimulus as seen were markedly reduced when positive displacement occurred with tracking off, with odds ratios of 0.16 [0.05, 0.50] (pd = 0.999) at locus 2, 0.18 [0.06, 0.49] (pd = 1.000) at locus 3, and 0.23 [0.05, 0.97] (pd = 0.977) at locus 4.

### Optimal criterion for suprathreshold (defect-mapping) microperimetry

Analysis of the operating characteristics derived from the fitted frequency-of-seeing functions identified a criterion intensity of 13 dB as nominally optimal for distinguishing “seeing” from “non-seeing” retinal loci in single-presentation suprathreshold testing (Figure 5). At this intensity, the Youden index reached 0.76 (95% credible interval [0.74, 0.79]). Notably, restricting the analysis to locus 1 (“seeing”) and locus 5 (“non-seeing”) yielded comparable results, indicating that the estimated optimal criterion was robust to potential confounding from peripapillary regions (Figure 5).

**Figure 5.**
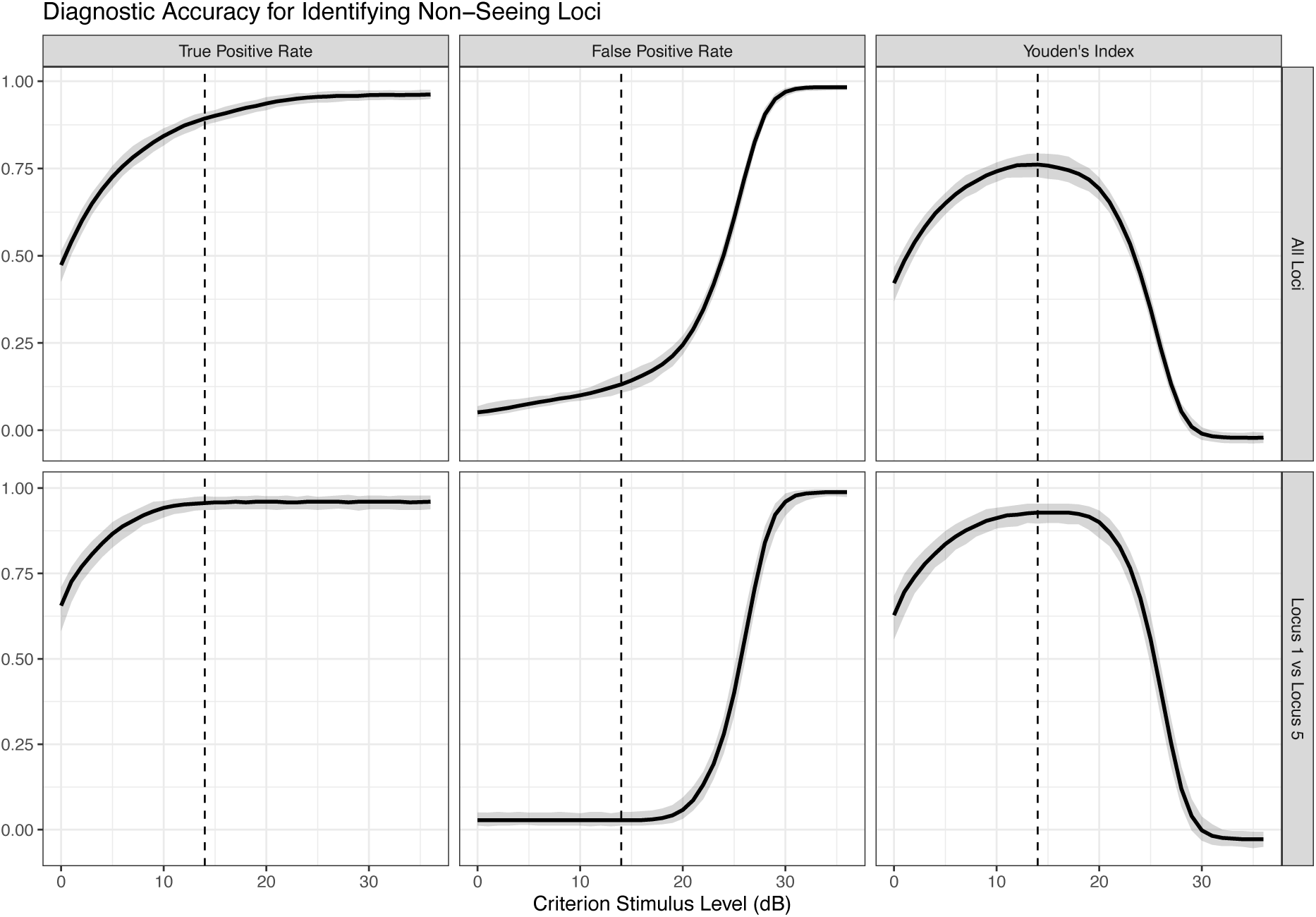
Determination of an optimal criterion for suprathreshold (defect-mapping) microperimetry. Using the fitted frequency-of-seeing curves, the figure illustrates how the choice of stimulus intensity affects the ability to identify “non-seeing” retinal loci with a single presentation. Curves depict the median (solid line) and 95% credible intervals (shaded bands) of the true-positive rate, false-positive rate, and the Youden index (right), plotted as a function of stimulus intensity. The vertical dashed line indicates the criterion intensity that maximizes the Youden index, representing the optimal trade-off between sensitivity and specificity. The lower row shows the same analysis including only locus 1 as “seeing” and locus 5 as “non-seeing” to minimize artifacts related to peripapillary atrophy, which could alter the psychometric function/threshold, especially at locus 3.

The 95% credible interval for the Youden index at 13 dB overlapped with that at 10 dB (Youden index 0.74 [0.72, 0.76]), suggesting similar overall discriminative performance across this range of criterion intensities. Criterion levels below approximately 10 dB were associated with a progressive decline in the Youden index, driven primarily by a reduction in the true-positive rate for identifying “non-seeing” loci. In contrast, criterion levels above approximately 16 dB showed a marked decrease in the Youden index due to an increasing false-positive rate, reflecting misclassification of “seeing” loci as “non-seeing”. Together, these findings indicate a relatively broad plateau of near-optimal performance between 10 and 15 dB, with diminishing diagnostic utility outside this range.

## DISCUSSION

Microperimetry has become a practical clinical-trial tool promising retinal-contingent stimulus delivery, especially for patients with fixation instability. In routine interpretation, non-responses within established scotomata (e.g., within GA) and steep sensitivity recovery just outside lesion borders are often taken as indirect validation that stimuli are delivered where intended.^2,6,8^ Yet, such “face validity” is an aggregate phenomenon; it does not resolve whether each individual presentation lands at its intended retinal location, particularly in regions with steep sensitivity gradients where sub-degree placement errors can change an outcome from “seen” to “not seen.” Here, we addressed this knowledge gap directly by prospectively sampling FoS functions with the MAIA device under controlled conditions, and by explicitly manipulating the fundus tracking state (tracking enabled vs disabled). The optic nerve head provided a natural, anatomically defined deep scotoma with a relatively sharp boundary,^12^ allowing us to evaluate stimulus delivery and psychometric behavior near the scotoma edge – precisely the context where tracking accuracy matters most for clinical monitoring.

Using the Open Perimetry Interface and a constant-stimuli FoS paradigm, we quantified how tracking changes the psychometric function as a surrogate for the tracking quality. Two patterns were evident. Firstly, thresholds in intact retina were largely unchanged by tracking, consistent with the expectation that small placement variability has a limited effect when sensitivity is high. Secondly, systematic tracking sharpened the psychometric transition (i.e., steepened the slope parameter), most importantly at loci closest to the scotoma boundary.

These observations are the psychophysical signature expected if tracking reduces trial-to-trial spatial jitter: the sensitivity estimates – especially in the “seeing retina” – as an aggregate phenomenon of many stimulus presentations change little, but the FoS steepen, meaning that sensitivity estimates become less uncertain. If tracking can further reduce effective stimulus-placement variability in this best-case scenario, the benefit in clinical cohorts, where fixation stability is frequently degraded by disease, is likely to be larger, particularly for endpoints that depend on classifying loci in junctional zones or at lesion borders.

The present data are directly relevant to suprathreshold microperimetry, also known as defect-mapping microperimetry. The defect-mapping paradigm, advanced by Wu and colleagues in the context of retinal diseases as an alternative to conventional staircase thresholding, replaces full threshold determination with a binary classification of function—“seeing” versus “non-seeing”—derived from responses to a single fixed-intensity stimulus delivered at each locus. ^8,12^ By avoiding threshold estimation, test time can be reallocated to higher spatial sampling density across the macula, thereby improving the ability to capture and follow complex scotoma topographies. ^8,12^

A central methodological issue for defect-mapping microperimetry is the choice of the criterion stimulus intensity. Two candidate criteria have been used in practice, 0 dB and 10 dB, but the two are not equivalent psychophysically. To date, three convergent lines of evidence have suggested that a criterion near 10 dB may be better suited for separating the “seeing” from the “non-seeing” retina than the maximum-intensity stimulus of 0 dB (equal to 318 cd/m^2^ on the mesopic MAIA scale). Firstly, 10 dB has been conceptualized as the lower bound of the “effective dynamic range,” recognizing that measurements at the physical floor are not stably reproduced on retest and that the probability of observing <0 dB is nontrivial even when true sensitivity is near the floor.^21^ Secondly, in a GA cohort, defect mapping with a 10 dB stimulus has demonstrated a closer association with imaging-derived atrophy metrics than defect mapping using 0 dB.^8^ Notably, responses to the 0 dB stimulus within areas of atrophy have been documented – either due to residual cone cells or stray light illumination.^11^ Thirdly, simulation analyses anchored in empirical microperimetry datasets indicate that repeatable <0 dB outcomes occur in only ∼60% of test–retest pairs even when a truly nonresponding location exists, whereas repeatable deep defects defined by ≤10 dB on both tests exceed 90% sensitivity while maintaining low false-positive rates.^11,13^ Together, these observations argue that 0 dB is an overly stringent operational definition of “non-seeing,” whereas 10 dB more closely tracks the reliably classifiable portion of deep sensitivity loss.

The present study adds a fourth and independent line of evidence by providing a psychometrically grounded criterion derived from FoS modeling. Specifically, FoS curves allow estimation of the stimulus intensity corresponding to a low detection probability (e.g., 5%), thereby defining a suprathreshold cut-off anchored to a measured response behavior rather than a heuristic convention. On this basis, a criterion stimulus level of 13 dB emerged as the nominally optimal separator between “seeing” and “non-seeing” loci. However, this estimate is anchored to normative sensitivity at “seeing” locations in healthy observers. In eyes with AMD, retina-wide photoreceptor degeneration and generalized mesopic sensitivity loss—typically on the order of 2–4 dB—are well documented.^22–24^ Accordingly, and given the broad plateau of near-equivalent performance observed for criterion intensities between 10 and 16 dB, selection of a 10 dB criterion for defect-mapping microperimetry represents a conservative and physiologically plausible choice that preserves discriminative performance while accommodating disease-related sensitivity loss.

### Limitations

First, the data were obtained from healthy volunteers, and the optic nerve head was used as a surrogate scotoma. Although this model offers the advantages of anatomical certainty and sharp boundaries, disease-related scotomata (e.g., GA) may differ in border complexity, residual function within lesions, and fixation behavior. Nonetheless, tracking-related effects were detectable even in this best-case cohort with relatively stable fixation, supporting the biological plausibility of a larger impact in patient populations where fixation is typically less stable and stimulus placement uncertainty is correspondingly higher. Second, device technology continues to evolve. The availability of newer hardware (e.g., MAIA3^25^) with improved tracking may alter the magnitude of effects observed here. However, this does not negate the primary conclusion that stimulus-placement variability measurably shapes psychometric behavior.

In summary, fundus tracking confers a modest yet consistent gain in measurement reliability, even under stable fixation, and is associated with steeper psychometric functions near scotoma boundaries. These data also provide a psychometric foundation for selecting the suprathreshold criterion used in defect-mapping paradigms. A cut-off near 13 dB (on the respective mesopic microperimetry scale) is supported as a reasonable and defensible criterion for separating “seeing” from “non-seeing” retina, thereby strengthening the scientific rationale of current defect-mapping microperimetry protocols as an efficient outcome measure in macular diseases

## Supporting information

Online Only Supplement

## Data Availability

All data generated in this study will be made publicly available in the Zenodo repository upon publication of this article.

## Disclosures

T. Lipsky: None;

C. Ehrenzeller: None;

G. Ansari: None;

K. Pfau: Daichii Sankyo (C), Inozyme (F), Heidelberg Engineering (R), Bayer (R), Roche (R);

W. Harmening: None;

Z. Wu: None;

M. Pfau: iCare (F), Inozyme (F), Roche (F)

