## Supplementary material for "How “Micro” is Microperimetry? Characterizing the Effect of Fundus Tracking on the Psychometric Function": Online Only Supplement

**Supplementary Figure 1. Test–retest reliability of psychometric function parameters.** The panels show Bland–Altman plots assessing test–retest reliability for the estimated psychometric function parameters: threshold m in dB (first row), slope parameter s in dB (second row), guess rate (third row), and lapse rate (fourth row). Points represent individual measurements (one point per test location, i.e., five per subject). The solid red horizontal lines indicate the mean bias and the upper and lower limits of agreement. Shaded red ribbons depict the corresponding 95% confidence intervals for the bias and limits of agreement.

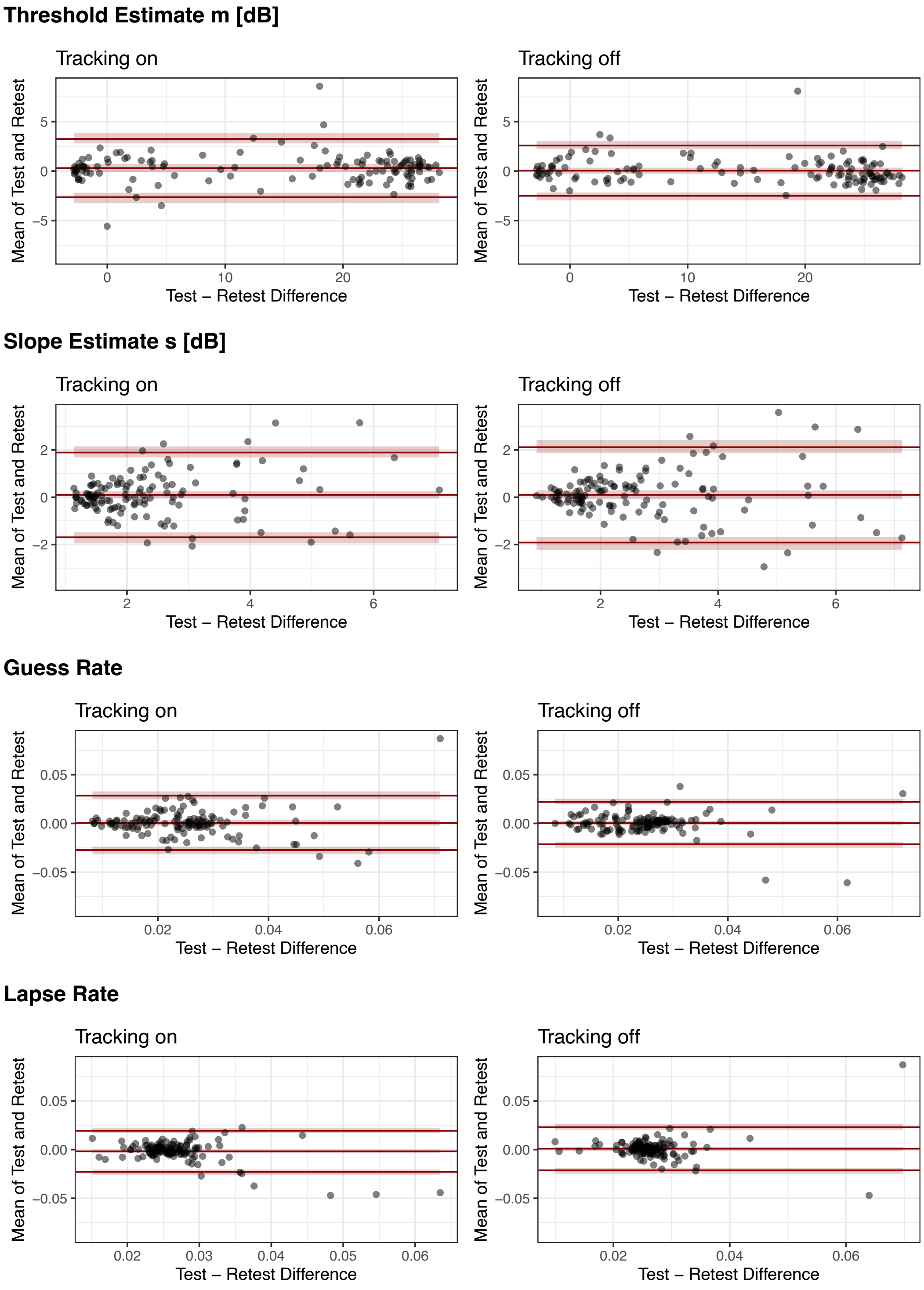

#### Supplementary Table S1. Test–Retest Reliability of Psychometric Function Parameters

| Parameter | Tracking | Bias [95% CI] | Lower LoA [95% CI] | Upper LoA [95% CI] |
|---|---|---|---|---|
| Threshold Estimate m [dB] | *on* | 0.30 [-0.10, 0.70] | -2.64 [-3.25, -2.19] | 3.25 [2.80, 3.86] |
| Threshold Estimate m [dB] | *off* | 0.04 [-0.27, 0.34] | -2.51 [-2.96, -2.16] | 2.58 [2.24, 3.04] |
| Slope Estimate s [dB] | *on* | 0.10 [-0.05, 0.25] | -1.69 [-1.95, -1.49] | 1.89 [1.69, 2.15] |
| Slope Estimate s [dB] | *off* | 0.10 [-0.09, 0.29] | -1.92 [-2.22, -1.68] | 2.12 [1.88, 2.42] |
| Lapse Rate | *on* | -0.00 [-0.00, -0.00] | -0.02 [-0.03, -0.02] | 0.02 [0.02, 0.02] |
| Lapse Rate | *off* | 0.00 [-0.00, 0.00] | -0.02 [-0.02, -0.02] | 0.02 [0.02, 0.03] |
| Guess Rate | *on* | 0.00 [-0.00, 0.00] | -0.03 [-0.03, -0.02] | 0.03 [0.03, 0.03] |
| Guess Rate | *off* | 0.00 [-0.00, 0.00] | -0.02 [-0.02, -0.02] | 0.02 [0.02, 0.03] |

**Supplementary Table S2. Comparison of Locus-Specific Effect of “Tracking On” Estimated from Joint and Separate Bayesian Models for Threshold and Slope Parameter**

| Outcome | Model | Locus 1<br>Median [95% CrI] (pd) | Locus 2<br>Median [95% CrI] (pd) | Locus 3<br>Median [95% CrI] (pd) | Locus 4<br>Median [95% CrI] (pd) | Locus 5<br>Median [95% CrI] (pd) |
|---|---|---|---|---|---|---|
| **Threshold** | Joint | −0.13 [−0.72, 0.44] (0.666) | 0.22 [−0.39, 0.81] (0.769) | −0.37 [−1.05, 0.31] (0.856) | −1.46 [−2.30, −0.62] (1.000) | −1.02 [−1.94, −0.08] (0.983) |
| | Separate | −0.14 [−0.72, 0.43] (0.685) | 0.21 [−0.37, 0.78] (0.767) | −0.39 [−1.06, 0.28] (0.878) | −1.46 [−2.32, −0.60] (1.000) | −1.03 [−1.96, −0.07] (0.983) |
| **Slope** | Joint | −0.14 [−0.29, 0.01] (0.967) | −0.27 [−0.43, −0.12] (1.000) | −0.22 [−0.40, −0.04] (0.988) | −0.02 [−0.24, 0.19] (0.558) | −0.18 [−0.38, 0.01] (0.965) |
| | Separate | −0.14 [−0.27, 0.00] (0.974) | −0.28 [−0.42, −0.13] (1.000) | −0.22 [−0.40, −0.04] (0.991) | −0.01 [−0.22, 0.20] (0.551) | −0.18 [−0.37, 0.01] (0.965) |

Values denote the posterior medians with 95% credible intervals [95% CrI] and probability of direction (pd).

The separate models treat threshold and slope parameter as independent outcomes, whereas the joint model analyzes them simultaneously, allowing partial pooling across outcomes via shared subject-level random effects. This can improve efficiency and stabilize estimates when outcomes are correlated. Residual correlations between threshold and slope parameter were not modeled because the two outcomes were fitted with outcome-specific inverse-variance weights, which preclude estimation of residual correlations in *brms*; however, cross-outcome associations at the subject level were retained through shared random effects. The results were highly similar between approaches, indicating weak cross-outcome correlation and robust conclusions.

### Supplementary Table S3. Frequentist Per-Locus Linear Mixed-Effects Models for the Effect of Tracking on Threshold

| | **Locus 1** | | | **Locus 2** | | | **Locus 3** | | | **Locus 4** | | | **Locus 5** | | |
|---|---|---|---|---|---|---|---|---|---|---|---|---|---|---|---|
| *Predictors* | *Estimates* | *CI* | *p* | *Estimates* | *CI* | *p* | *Estimates* | *CI* | *p* | *Estimates* | *CI* | *p* | *Estimates* | *CI* | *p* |
| (Intercept) | 25.77 | 25.14 – 26.40 | **<0.001** | 23.13 | 21.11 – 25.15 | **<0.001** | 17.97 | 14.75 – 21.19 | **<0.001** | 6.46 | 3.68 – 9.23 | **<0.001** | 0.56 | -0.87 – 1.99 | 0.440 |
| Tracking [on] | -0.29 | -0.67 – 0.09 | 0.132 | 0.20 | -0.32 – 0.71 | 0.445 | 0.03 | -0.88 – 0.93 | 0.950 | -1.18 | -2.24 – -0.11 | **0.030** | -1.22 | -2.06 – -0.38 | **0.005** |
| **Random Effects** | | | | | | | | | | | | | | | |
| $\sigma^2$ | 3.83 | | | 3.38 | | | 1.65 | | | 2.01 | | | 0.93 | | |
| $\tau_{00}$ | 2.27 $_{subject}$ | | | 25.58 $_{subject}$ | | | 65.47 $_{subject}$ | | | 48.20 $_{subject}$ | | | 12.60 $_{subject}$ | | |
| $\tau_{11}$ | 0.43 $_{subject.trackingon}$ | | | 1.14 $_{subject.trackingon}$ | | | 4.56 $_{subject.trackingon}$ | | | 5.95 $_{subject.trackingon}$ | | | 3.75 $_{subject.trackingon}$ | | |
| $\rho_{01}$ | 0.22 $_{subject}$ | | | -0.53 $_{subject}$ | | | -0.42 $_{subject}$ | | | -0.23 $_{subject}$ | | | -0.55 $_{subject}$ | | |
| ICC | 0.41 | | | 0.87 | | | 0.97 | | | 0.96 | | | 0.92 | | |
| N | 25 $_{subject}$ | | | 25 $_{subject}$ | | | 25 $_{subject}$ | | | 25 $_{subject}$ | | | 25 $_{subject}$ | | |
| Observations | 100 | | | 100 | | | 100 | | | 100 | | | 100 | | |
| Marginal $R^2$ / Conditional $R^2$ | 0.003 / 0.416 | | | 0.000 / 0.873 | | | 0.000 / 0.973 | | | 0.007 / 0.960 | | | 0.031 / 0.923 | | |

### Supplementary Table S4. Frequentist Per-Locus Linear Mixed-Effects Models for the Effect of Tracking on Slope Parameter

| | **Locus 1** | | | **Locus 2** | | | **Locus 3** | | | **Locus 4** | | | **Locus 5** | | |
|---|---|---|---|---|---|---|---|---|---|---|---|---|---|---|---|
| *Predictors* | *Estimates* | *CI* | *p* | *Estimates* | *CI* | *p* | *Estimates* | *CI* | *p* | *Estimates* | *CI* | *p* | *Estimates* | *CI* | *p* |
| (Intercept) | 1.63 | 1.48 – 1.79 | **<0.001** | 1.96 | 1.74 – 2.18 | **<0.001** | 2.92 | 2.38 – 3.47 | **<0.001** | 3.13 | 2.59 – 3.66 | **<0.001** | 2.36 | 1.95 – 2.76 | **<0.001** |
| Tracking [on] | -0.14 | -0.27 – -0.02 | **0.022** | -0.30 | -0.50 – -0.10 | **0.004** | -0.41 | -0.82 – 0.01 | 0.057 | -0.05 | -0.67 – 0.56 | 0.863 | -0.38 | -0.85 – 0.09 | 0.112 |
| **Random Effects** | | | | | | | | | | | | | | | |
| $\sigma^2$ | 0.50 | | | 0.99 | | | 0.65 | | | 0.70 | | | 0.70 | | |
| $\tau_{00}$ | 0.11 subject | | | 0.21 subject | | | 1.66 subject | | | 1.42 subject | | | 0.80 subject | | |
| $\tau_{11}$ | 0.03 subject.trackingon | | | 0.08 subject.trackingon | | | 0.75 subject.trackingon | | | 1.66 subject.trackingon | | | 0.95 subject.trackingon | | |
| $\rho_{01}$ | -0.57 subject | | | -0.80 subject | | | -0.68 subject | | | -0.59 subject | | | -0.84 subject | | |
| ICC | 0.16 | | | 0.13 | | | 0.66 | | | 0.66 | | | 0.44 | | |
| N | 25 subject | | | 25 subject | | | 25 subject | | | 25 subject | | | 25 subject | | |
| Observations | 100 | | | 100 | | | 100 | | | 100 | | | 100 | | |
| Marginal $R^2$/ Conditional $R^2$ | 0.009 / 0.166 | | | 0.020 / 0.144 | | | 0.021 / 0.670 | | | 0.000 / 0.657 | | | 0.028 / 0.454 | | |

**Supplementary Table S5. Bayesian Joint Model Estimates of Odds Ratios for False-Positive Responses Across Loci**

Odds ratios (OR), 95% credible intervals (CrI), and probability of direction (pd) for the effects of negative fixation shifts (shifts of the stimulus toward the "seeing" retina), tracking status, and their interaction on the likelihood of false-positive responses, estimated from a joint Bayesian logistic regression model across all loci.

| Predictor | Locus 1 OR (95% CrI) | pd | Locus 2 OR (95% CrI) | pd | Locus 3 OR (95% CrI) | pd | Locus 4 OR (95% CrI) | pd | Locus 5 OR (95% CrI) | pd |
|---|---|---|---|---|---|---|---|---|---|---|
| (Intercept) | 0.02 [0.01, 0.03] | 1 | 0.01 [0.01, 0.02] | 1 | 0.01 [0.00, 0.03] | 1 | 0.04 [0.02, 0.07] | 1 | 0.03 [0.02, 0.05] | 1 |
| Negative Fixation Shift | 0.96 [0.58, 1.62] | 0.557 | 1.99 [1.00, 4.23] | 0.975 | 1.57 [0.65, 4.12] | 0.84 | 1.35 [0.65, 2.81] | 0.785 | 1.69 [1.06, 2.74] | 0.985 |
| Tracking (Off vs On) | 2.62 [1.64, 4.34] | 1 | 3.21 [1.56, 6.90] | 0.999 | 2.93 [1.15, 7.78] | 0.987 | 2.93 [1.46, 6.01] | 1 | 2.15 [1.32, 3.50] | 0.999 |
| Negative Fixation Shift × Tracking (Off) | 1.27 [0.69, 2.24] | 0.795 | 1.20 [0.51, 2.71] | 0.676 | 3.62 [1.24, 10.69] | 0.992 | 2.14 [0.87, 5.12] | 0.956 | 2.90 [1.63, 5.26] | 1 |

### Supplementary Table S6. Frequentist Mixed-Effects Logistic Regression Models for False-Positive Responses by Locus

Odds ratios (OR), 95% confidence intervals (CI), and p-values from per-locus generalized linear mixed-effects models assessing the effects of negative fixation shifts (shifts of the stimulus toward the “seeing” retina), tracking, and their interaction on false-positive responses.

| | **Locus 1** | | | **Locus 2** | | | **Locus 3** | | | **Locus 4** | | | **Locus 5** | | |
|---|---|---|---|---|---|---|---|---|---|---|---|---|---|---|---|
| *Predictors* | *Odds Ratios* | *CI* | *p* | *Odds Ratios* | *CI* | *p* | *Odds Ratios* | *CI* | *p* | *Odds Ratios* | *CI* | *p* | *Odds Ratios* | *CI* | *p* |
| (Intercept) | 0.02 | 0.01 – 0.03 | **<0.001** | 0.01 | 0.01 – 0.03 | **<0.001** | 0.01 | 0.00 – 0.03 | **<0.001** | 0.04 | 0.01 – 0.10 | **<0.001** | 0.03 | 0.01 – 0.05 | **<0.001** |
| Negative Fixation Shift | 1.46 | 0.80 – 2.67 | 0.223 | 2.37 | 1.07 – 5.25 | **0.034** | 1.29 | 0.45 – 3.70 | 0.641 | 1.31 | 0.60 – 2.90 | 0.499 | 1.38 | 0.83 – 2.29 | 0.209 |
| Tracking (Off vs On) | 2.73 | 1.58 – 4.72 | **<0.001** | 3.45 | 1.52 – 7.84 | **0.003** | 2.91 | 0.99 – 8.56 | 0.053 | 3.18 | 1.56 – 6.50 | **0.001** | 2.20 | 1.33 – 3.63 | **0.002** |
| Negative Fixation Shift × Tracking (Off) | 1.16 | 0.58 – 2.34 | 0.672 | 1.03 | 0.41 – 2.56 | 0.958 | 4.28 | 1.26 – 14.57 | **0.020** | 2.20 | 0.85 – 5.69 | 0.106 | 3.38 | 1.84 – 6.23 | **<0.001** |
| **Random Effects** | | | | | | | | | | | | | | | |
| $\sigma^2$ | 3.29 | | | 3.29 | | | 3.29 | | | 3.29 | | | 3.29 | | |
| $\tau_{00}$ | 0.09 $_{subject}$ | | | 0.32 $_{subject}$ | | | 1.03 $_{subject}$ | | | 1.48 $_{subject}$ | | | 1.37 $_{subject}$ | | |
| ICC | 0.03 | | | 0.09 | | | 0.24 | | | 0.31 | | | 0.29 | | |
| N | 25 $_{subject}$ | | | 18 $_{subject}$ | | | 12 $_{subject}$ | | | 9 $_{subject}$ | | | 20 $_{subject}$ | | |
| Observations | 4192 | | | 2722 | | | 1875 | | | 1146 | | | 3219 | | |
| Marginal $R^2$ / Conditional $R^2$ | 0.097 / 0.120 | | | 0.144 / 0.219 | | | 0.241 / 0.421 | | | 0.143 / 0.409 | | | 0.152 / 0.402 | | |

**Supplementary Table S7. Bayesian Joint Model Estimates of Odds Ratios for False-Negative Responses Across Loci**

Odds ratios (OR), 95% credible intervals (CrI), and probability of direction (pd) for the effects of positive fixation shifts (shifts of the stimulus toward the optic nerve head), tracking status, and their interaction on the likelihood of false-negative responses, estimated from a joint Bayesian logistic regression model across all loci.

| Predictor | Locus 1 OR [95% CrI] | pd | Locus 2 OR [95% CrI] | pd | Locus 3 OR [95% CrI] | pd | Locus 4 OR [95% CrI] | pd |
|---|---|---|---|---|---|---|---|---|
| (Intercept) | 18.58 [10.21, 35.08] | 1.000 | 21.25 [11.31, 44.96] | 1.000 | 15.18 [7.95, 31.59] | 1.000 | 14.77 [5.69, 43.06] | 1.000 |
| Positive Fixation Shift | 1.10 [0.51, 2.36] | 0.585 | 1.55 [0.60, 4.04] | 0.815 | 1.22 [0.54, 2.92] | 0.678 | 1.59 [0.47, 5.58] | 0.772 |
| Tracking (Off vs On) | 0.85 [0.41, 1.77] | 0.667 | 0.86 [0.35, 2.05] | 0.626 | 0.52 [0.26, 1.05] | 0.965 | 1.09 [0.35, 3.27] | 0.559 |
| Positive Fixation Shift × Tracking (Off) | 0.61 [0.22, 1.61] | 0.843 | 0.16 [0.05, 0.50] | 0.999 | 0.18 [0.06, 0.49] | 1.000 | 0.23 [0.05, 0.97] | 0.977 |

### Supplementary Table S8. Frequentist Mixed-Effects Logistic Regression Models for False-Negative Responses by Locus

Odds ratios (OR), 95% confidence intervals (CI), and p-values from per-locus generalized linear mixed-effects models assessing the effects of positive fixation shifts (shifts of the stimulus toward the optic nerve head), tracking, and their interaction on false-negative responses.

| | Locus 1 | | | Locus 2 | | | Locus 3 | | | Locus 4 | | |
|---|---|---|---|---|---|---|---|---|---|---|---|---|
| *Predictors* | *Odds Ratios* | *CI* | *p* | *Odds Ratios* | *CI* | *p* | *Odds Ratios* | *CI* | *p* | *Odds Ratios* | *CI* | *p* |
| (Intercept) | 25.67 | 12.67 – 52.00 | **<0.001** | 21.10 | 11.19 – 39.79 | **<0.001** | 15.83 | 7.32 – 34.26 | **<0.001** | 10.80 | 4.32 – 27.00 | **<0.001** |
| Positive Fixation Shift | 0.67 | 0.26 – 1.76 | 0.419 | 1.61 | 0.58 – 4.51 | 0.364 | 1.26 | 0.49 – 3.24 | 0.626 | 2.19 | 0.50 – 9.57 | 0.297 |
| Tracking (Off vs On) | 0.62 | 0.25 – 1.55 | 0.305 | 1.12 | 0.42 – 3.02 | 0.817 | 0.56 | 0.26 – 1.17 | 0.124 | 1.44 | 0.40 – 5.23 | 0.576 |
| Positive Fixation Shift × Tracking (Off) | 1.25 | 0.34 – 4.59 | 0.741 | 0.11 | 0.03 – 0.42 | **0.001** | 0.16 | 0.05 – 0.48 | **0.001** | 0.14 | 0.02 – 0.92 | **0.040** |
| **Random Effects** | | | | | | | | | | | | |
| $\sigma^2$ | 3.29 | | | 3.29 | | | 3.29 | | | 3.29 | | |
| $\tau_{00}$ | 0.10 $_{subject}$ | | | 0.00 $_{subject}$ | | | 0.31 $_{subject}$ | | | 0.00 $_{subject}$ | | |
| ICC | 0.03 | | | | | | 0.09 | | | | | |
| N | 9 $_{subject}$ | | | 10 $_{subject}$ | | | 5 $_{subject}$ | | | 2 $_{subject}$ | | |
| Observations | 722 | | | 788 | | | 572 | | | 275 | | |
| Marginal $R^2$ / Conditional $R^2$ | 0.018 / 0.047 | | | 0.157 / NA | | | 0.159 / 0.231 | | | 0.088 / NA | | |
